# Variants of SARS-COV-2 and the Death Toll

**DOI:** 10.1101/2021.07.08.21260081

**Authors:** Takesi Saito

## Abstract

New variants of SARS-COV-2 have been found in various countries. Especially, the UK has been attacked by India’s Delta Plus, and the spread of infection has been very rapid, since it is extremely infectious. Fortunately, however, the number of deaths has been stayed flat, where deaths are reported to be those who are not yet received a shot of COVID-19 vaccine. In this short not, we would like to consider why the number of deaths is so small, compared with high cases, around the infection peak, when the basic reproduction number is very large.

## Formulation and Discussion

We consider this problem by means of the SIR model [1], [2∼13] in the theory of infection. Equations of the SIR model are given by

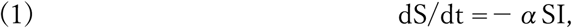

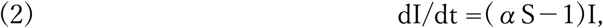

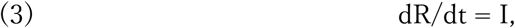

where S, I and R are numbers for susceptibles, infectives and removed, respectively, and *α* is the basic reproduction number. The removed ratio c is set to be c=1 for simplicity. We normalize S, I and R as

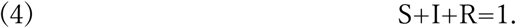

At the peak of infection t=T, following formulas are well known to hold

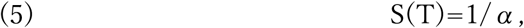

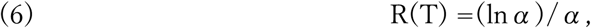

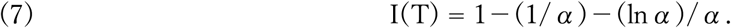

Curves of R(T) and I(T) against α are drawn in Fig. 1. From Fig. 1 we see that the larger I(T), then the smaller R(T) for α ≥ 3.51 at the peak. That is to say, at the peak the removed number decreases as the infection number increases for α ≥ 3.51. For *α*=5, as an example of higher cases, we see that I(T) = 0.48 is larger than R(T) = 0.32. Conversely, for α = 2.5, as an example of lower cases, we get I(T) = 0.26 is smaller than R(T)=0.37. Once having α = 5 and 2.5, we can draw curves for S, I and R against the time t by means of Excell in Figs. 2 and 3, respectively. In Fig.2 one can see that the peak value I(T) is correctly lager than R(T), while in Fig.3 this relationship is reversely.

**Fig1.**
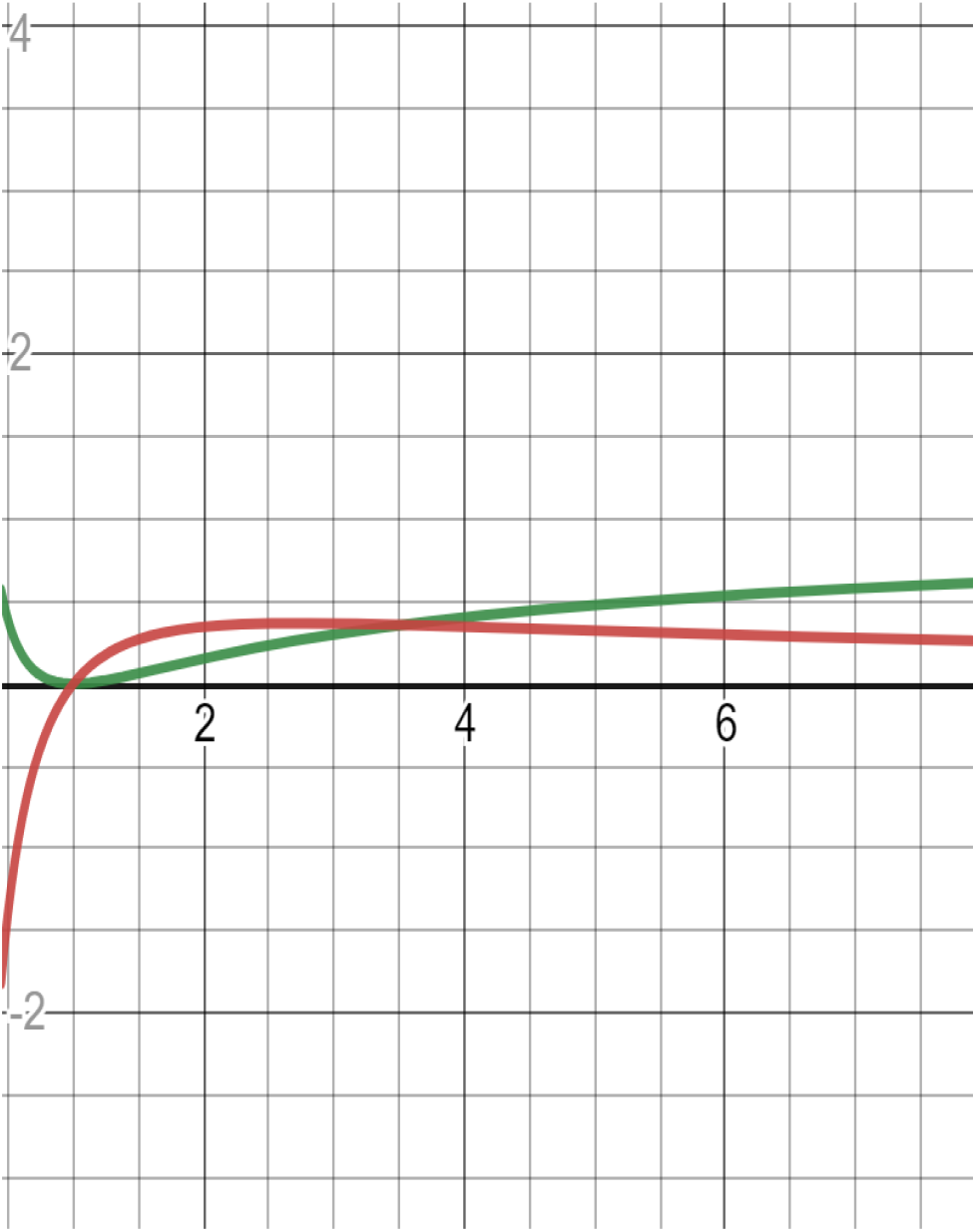
The red curve is for R(T)=(lnα)/α. The blue curve is for I(T) = 1 − (1/α)−(lnα)/α. The horizontal line corresponds to α ≥ 1. The crossing point for both curves is α = 3.51.

**Fig. 2.**
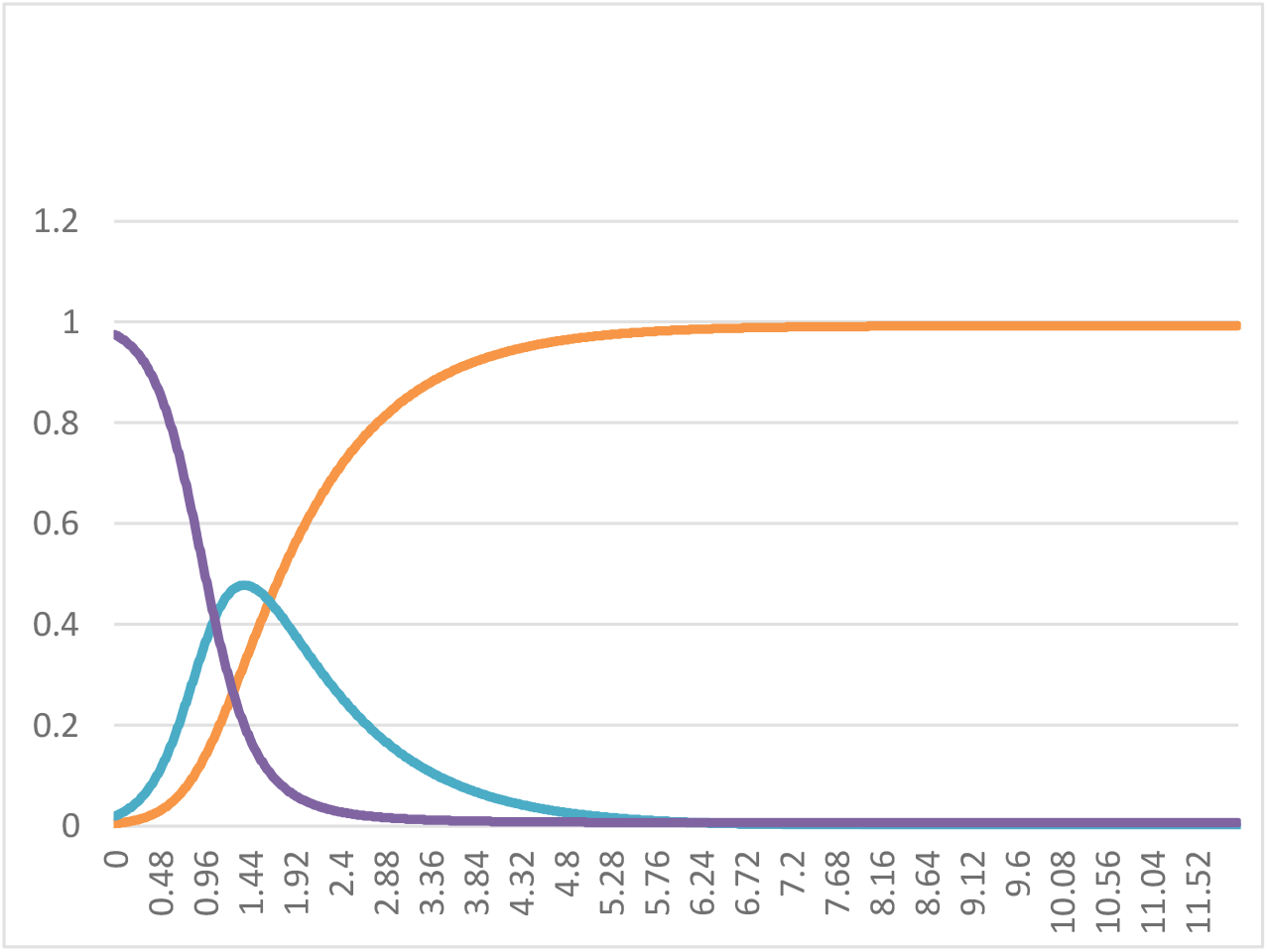
Curves for S(deep-blue), I(blue) and R(red) with α = 5, c = 1, where S, I and R are numbers for susceptibles, infectives and removed, respectively, and *α* is the basic reproduction number, c the removed ratio. The vertical line corresponds to S, I and R. The horizontal line corresponds to a time.

**Fig. 3.**
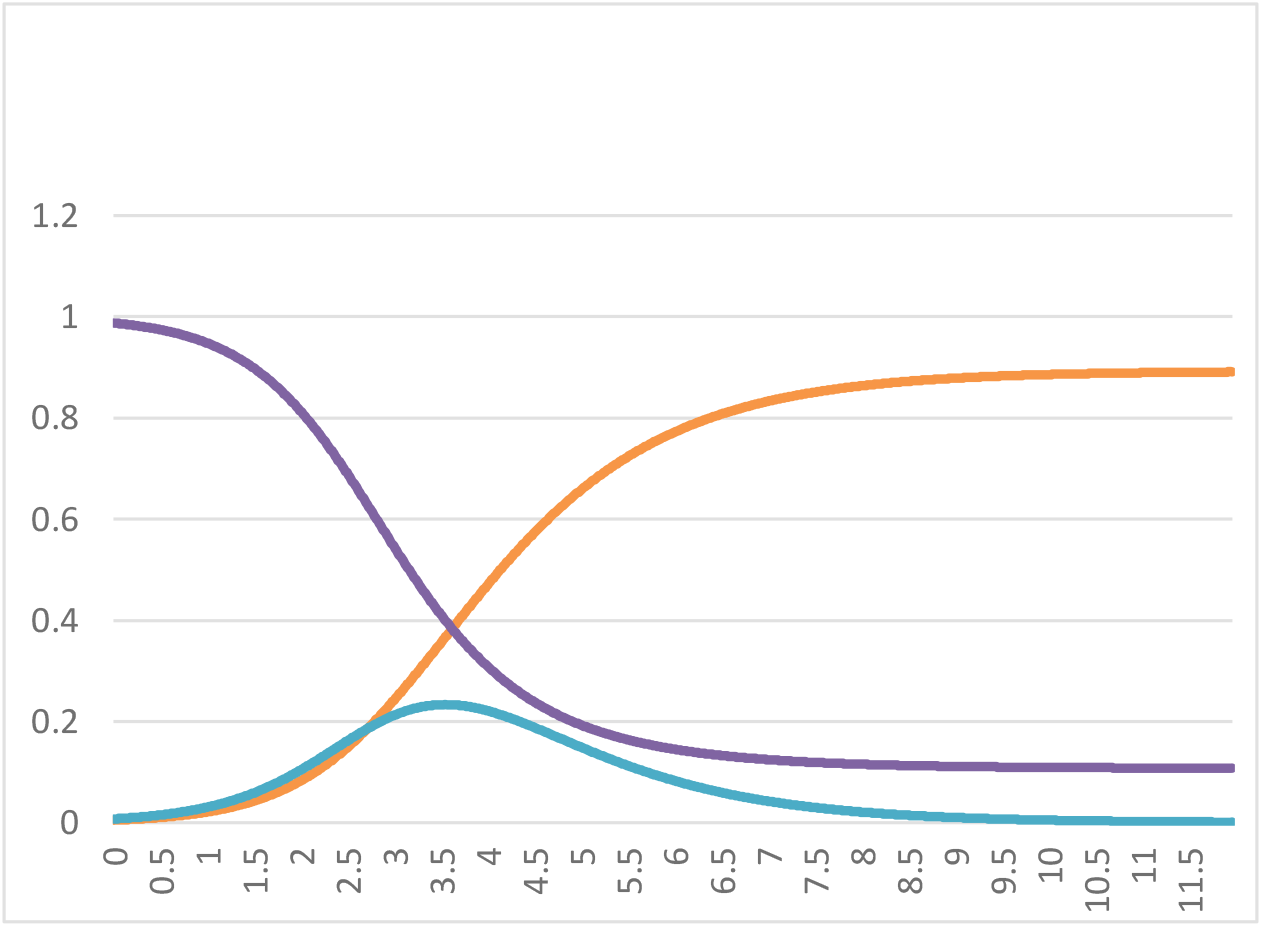
Curves for S(deep-blue), I(blue) and R(red) with α = 2. 5, c = 1.

Let us define the morality ratio by λ = D(t)/R(t), where D(t) is the accumulated number of deaths at t. We assume λ to be approximately constant [14]. Hence D(t) is proportional to R(t). Our aim is to consider the reason why the number of deaths is so small as compared with that of high cases at the peak, t = T, when *α* is very large. From above discussions, we conclude that as α gets bigger, I(T) increases, then R(T) decreases inversely, hence D(T)= λR(T) does also so.

After passing the peak, we get R(t) > *I*(*t*), as is seen in Fig. 2 with α = 5. However, from the formula D(∞) = λR(∞) ≅ λ, there is a possibility that the accumulate number D(∞) at the end may be still small, when λ is very small. Otherwise, of course, D(∞) may be large.

## Conclusion

In conclusion, we succeeded to explain why the number of deaths is so small, compared with high cases, at the infectious peak, when the basic reproduction α is very large. This essentially comes from Eqs.(6) and (7) derived from the SIR equation. The larger the basic reproduction number α, the larger the infectious number I(T). The larger the infectious number I(T), then the smaller the removed number R(T), hence the smaller the death toll D(T), around the infectious peak.

After passing the peak, we get R(t) > *I*(*t*), as is seen in Fig. 2 with α = 5. However, from the formula D(∞) = λR(∞) ≅ λ, there is a possibility that the accumulate number D(∞) at the end may be still small, when λ is very small. Otherwise, of course, D(∞) may be large.

## Data Availability

The availability of all data is OK.

https://toyokeizai.net/sp/visual/tko/covid19

## Acknowledgment

The author would like to express his deep gratitude to K. Shigemoto for valuable discussions.

